# Expression of mechano-growth factor (MGF) in refractory overactive bladder

**DOI:** 10.1101/2023.12.08.23299594

**Authors:** E Spiritosanto, B Lemmon, F Mohamedi-Yousufi, HA Munasinghe, A Mahmood, R Bray, FE Mackenzie, NJ Hill, E Cortes

**Author notes:** Denotes equal contribution as first authors (E Spiritosanto, B Lemmon). Equal contribution as senior authors (NJ Hill, E Cortes). Corresponding author: Natasha Hill, School of Biomedical Science, Faculty of Health, John Bull Building, Research Way, Plymouth Science Park, Plymouth PL6 8BU.

## Abstract

Overactive bladder (OAB) is a urological symptom complex defined by urinary urgency. It can have a devastating impact on an individual’s quality of life and leads to significant financial cost. Insulin-like growth factor 1 (IGF-1) is a protein hormone involved in a broad range of processes including cell proliferation and differentiation. IGF-1 is also regulated through alternative splicing and the alternative *IGF-1Ec* transcript encodes the proteolytically-derived MGF peptide. The role of MGF in skeletal muscle is well established but its role in smooth muscle, such as the detrusor muscle of the bladder, has been little explored. The aim of this study was to explore the expression of MGF in bladder biopsies from patients with refractory OAB and age-matched controls.

We show using immunohistochemistry that MGF is widely expressed in bladder tissue. Quantification of MGF expression by western blot showed MGF expression is similar in OAB and control bladder tissue. RT-qPCR analysis showed a significant decrease in the smooth muscle marker *DES* in OAB biopsies, potentially reflecting smooth muscle loss. While we did not observe any correlation of MGF expression and clinical measurements, our *in vitro* data suggests that MGF induces cell proliferation in primary bladder cultures, suggesting a potential regenerative role. Together our data therefore suggest that MGF is unlikely to be involved in the development of OAB pathology but may have therapeutic potential in OAB patients as an inducer of bladder smooth muscle proliferation to regenerate lost or dysfunctional bladder smooth muscle.

## Introduction

Overactive bladder (OAB) is a urological symptom complex defined by urinary urgency with or without urgency incontinence, usually accompanied by daytime frequency and nocturia, in the absence of proven urinary infection or other pathology^1^. The prevalence of OAB varies across studies but increases with age and disproportionately affects the female population, with up to 43% of women over forty having symptoms^2^. OAB can have a devastating impact on an individual’s quality of life and leads to significant direct and indirect financial cost^3^.

OAB is a symptomatic diagnosis based on taking a focused patient history with the aid of validated questionnaires. First line treatment includes lifestyle modifications, pelvic floor exercises, fluid management, and bladder retraining. Medical therapy is second line treatment, where receptors in the detrusor muscle are targeted to induce smooth muscle relaxation^4^. In the absence of improvement following conservative and medical management, a patient is described as having refractory OAB. Refractory OAB can be treated using techniques that encourage neuromodulation such as Botulinum toxin A (Botox) injection into the bladder muscle. Reliance on symptom analysis alone can lead to misdiagnosis and delays in accessing treatment. A more holistic approach that considers the physiological nature of OAB is needed.

Insulin-like growth factor 1 (IGF-1) is a protein hormone involved in a broad range of processes including cell growth and survival, proliferation, differentiation and tissue metabolism^5^. One of the mechanisms through which circulating IGF-1 is regulated and exerts its function is through alternative splicing, a process that allows the production of multiple mRNA transcripts encoding distinct proteins dependent on the tissue, developmental stage or external stimuli^6,7^. The precursor protein products of the different IGF-1 transcripts are cleaved to generate the mature IGF-1 protein and an E-peptide. The IGF-1 protein encoded by the different transcripts is identical but the E-peptides generated are distinct. The E-peptide generated by the *IGF-1Ec* transcript is commonly known as mechano-growth factor, or MGF. MGF is upregulated following mechanical stretch, injury and/or electrical stimulation of skeletal muscle *in vivo*^8,9^ and promotes skeletal muscle proliferation *in vitro*^10–12^. Changes in the regulation of MGF expression seen with age are associated with reduced ability to respond to mechanical overload^10,13,14^. MGF has also been shown to play a role in other tissues, including the brain^15^. Li *et al* showed that Transforming Growth Factor-β (TGF-β) mediated-IGF-1 production and smooth muscle cell hyperplasia is accompanied by MGF upregulation and peptide expression, with consequent hypertrophy of smooth muscle cells^16^. However, little is otherwise known about MGF in smooth muscle.

MGF is widely expressed in the wall of the human bladder^17^. We therefore hypothesised that reduced MGF expression may play a role in the development of OAB. In the present study, we examined the expression of MGF in human bladder biopsies of patients with refractory OAB. We evaluated the correlation between MGF expression and symptom severity at diagnosis using both validated symptom questionnaires and urodynamic parameters and tested the effect of MGF on bladder cell proliferation *in vitro*.

## Materials and Methods

### Patient selection

The study was granted ethical approval by the South-East London Research Ethics Committee (A5-2: 10/H0805/51, IRAS number 474470, 8^th^ Feb 2010). The chief investigator (EC) conducted the first phase of the study in collaboration with Kings College London between 2010-13. The second phase of the study was conducted in collaboration with Kingston University London between 2016-19. All experiments were performed in accordance with the declaration of Helsinki, and written informed consent was obtained from all study participants, and all institutional forms have been stored appropriately.

Patients included in the study group had a clinical diagnosis of refractory OAB (rOAB) and failed symptom improvement with bladder retraining and oral treatment with an anticholinergic drug, a beta-3 agonist (mirabegron), or both. All these women underwent urodynamic investigation as per NICE guidance. During urodynamic testing, first desire to void (FDV), normal sensation to void (NS), and Maximum Cystometric Capacity (MCC) were recorded for all participants. They were then discussed in our local multidisciplinary team (MDT) meeting at Kingston hospital and offered third line treatment for OAB with intra-vesical Botulinum Toxin A (Botox) injections. Prior to Botox treatment, women were consented by our two Urogynaecology Consultants and bladder biopsies were taken with rigid cystoscopy using cold cup biopsy forceps. Diathermy to the biopsy site was carried out to ensure haemostasis prior to proceeding with injections of either 100iu or 200iu of Botox. The control group was recruited from patients undergoing other gynaecological surgery without any lower urinary tract symptoms (LUTS). We asked women to complete two validated self-assessment screening questionnaires for OAB: OAB V-8 (Overactive Bladder Awareness Tool) and BSAQ (Bladder Control Self-Assessment Questionnaire) for mixed incontinence.

All bladder biopsies were immediately placed into an RNAlater buffer (Merck Life Sciences). For processing, samples were placed on absorbent paper, weighed and then homogenised using a TissueLyser (Qiagen) with 5 mm beads for 15 minutes at 20 Hz. Equal parts of the homogenate were taken for protein and RNA extraction using the PARIS extraction kit (ThermoFisher) according to the manufacturer’s protocol.

**Full details of laboratory techniques, including immunohistochemistry, western blot, RT-qPCR and proliferation assays are available as Supplementary Materials.**

### Statistical analysis

Statistical analysis was performed using RStudio, or MS Excel where indicated. Variables were tested for normality using the Shapiro-Wilk test, and significance levels greater than 0.05 were assumed to be normally distributed. Where data were not normally distributed or sample sizes were small the Mann–Whitney U test was used for two-sample comparisons. One-way analysis of variance (ANOVA) followed by the post hoc Tukey’s test was used to compare the means of more than two samples. Non-parametric Spearman’s rank-order correlation was used to assess the relationship between variables. Where tied ranks were present approximate p-values were calculated. To analyse the distribution of groups a χ^2^ test was performed in MS Excel; p-values ≤ 0.05 were considered statistically significant. Graphs were generated in either Excel or RStudio using ggplot.

## Results

### Study population

Our study population included 33 women with refractory OAB (rOAB) and detrusor overactivity on urodynamic testing, and 6 women without lower urinary tract symptoms as our controls (Table 1). Control patients had a mean age of 58.8 +/- 4.1 years and an average parity of two. Our study group had a mean age of 60.1 +/- 2.8 years and an average parity of two.

**Table 1.** Clinical characteristics of patient cohort. Clinical OAB parameters obtained with the urodynamic test *i.e ‘*1^st^‘ and ‘2^nd^‘ sensation and Maximum Cystometric Capacity (MCC). Self-assessment questionnaire scores (BSAQ and OABV8) were also recorded, although this information was only available for 31/40 patients. NA indicates data not available.

| Type | Age group | Ethnicity | Parity | 1st sensation | 2nd sensation | MCC | DO | Incontinence | BSAQ | OABV8 |
| --- | --- | --- | --- | --- | --- | --- | --- | --- | --- | --- |
| Control | ≤50 | White | 2 |  |  |  |  |  |  |  |
| Control | ≤50 | White | 2 | NA | NA | NA | NA |  | 3 | 5 |
| Control | >50 | White | 3 | NA | NA | NA | NA | no |  |  |
| Control | >50 | white | 2 | NA | NA | NA | NA | no | NA | NA |
| Control | >50 | White |  | NA | NA | NA | NA | no | 6 | 0 |
| Control | >50 | White |  |  |  |  |  |  | 2 | 2 |
| OAB | >50 | White | 1 | 120 | 210 | 410 | terminal | yes | 21 | 30 |
| OAB | >50 | Asian | 3 | 157 | 460 | 480 | yes | yes | 22 | 35 |
| OAB | >50 | Arab | 4 |  |  |  | yes |  |  |  |
| OAB | >50 | White |  | 85 | 340 | 450 | yes | yes |  |  |
| OAB | ≤50 | White | 0 | 130 | 200 | 240 | no |  | 17 | 21 |
| OAB | >50 | Other | twins | 125 | 275 | 518 | yes | yes | 22 | 28 |
| OAB | ≤50 | White | twins +1 | 158 | 300 | 340 | yes | yes | 24 | 39 |
| OAB | NA |  |  |  |  |  |  |  |  |  |
| OAB | >50 | White |  | 154 | 260 | 390 | no | yes | 19 | 18 |
| OAB | >50 | White | 4 | 173 | 324 | 415 | no | yes | 24 | 35 |
| OAB | >50 | White | 1 | 80 |  |  | yes | yes | 20 | 33 |
| OAB | ≤50 | White | 1 | 107 | 163 | 403 | yes | yes | 18 | 25 |
| OAB | ≤50 | Black | 4 | 95 | 205 | 359 | yes | yes | 22 | 23 |
| OAB | ≤50 | Other | 2 | NA | NA | NA | NA | yes | 23 | 38 |
| OAB | >50 | White | 3 | NA | NA | NA | NA |  | 13 | 13 |
| OAB | >50 | White |  | 100 | 300 | 429 | no | yes | 17 | 17 |
| OAB | ≤50 | Asian | 2 | NA | NA | NA | NA | no | 12 | 20 |
| OAB | >50 | White |  | 302 | 440 | 468 | yes | no | 23 | 35 |
| OAB | >50 | White | 1 | NA | NA | NA | NA | no | 15 | 32 |
| OAB | >50 | White | 2 | NA | NA | NA | NA | no | 12 | 10 |
| OAB | ≤50 | White | 1 | 56 | 311 | 401 | yes | yes | 20 | 33 |
| OAB | >50 | White | 1 | 120 | 233 | 500 | yes | no | 22 | 26 |
| OAB | >50 | White | 1 | 206 | 421 | 497 | yes | yes | 20 | 24 |
| OAB | >50 | white | 2 | 160 | 279 | 388 | yes | yes | NA | NA |
| OAB | >50 | Asian | 1 | 200 | 311 | 500 | yes | no | 18 | 33 |
| OAB | >50 | White | 2 | 83 | 250 | 357 | yes | no | NA | NA |
| OAB | ≤50 | White | 2 | 296 | 488 | 511 | no | no | 17 | 25 |
| OAB | >50 | White |  | 180 | 369 | 417 | yes | no | 7 | 16 |
| OAB | >50 | White |  | NA | NA | NA | NA | yes | NA | NA |
| OAB | >50 | White |  | 200 | 270 | 384 | yes | yes | 16 | 31 |
| OAB | >50 | White |  | 136 | 228 | 0 | yes | yes | 24 | 37 |
| OAB | >50 | White |  | 110 | 303 | 387 | yes | yes | 15 | 28 |
| OAB | >50 | White |  | 164 | 376 | 507 | yes | yes | 18 | 26 |

### Expression of the smooth muscle marker *DES* is reduced in OAB biopsies

The bladder consists of three main layers: urothelium, sub-mucosal lamina propria containing myofibroblast-like cells, and detrusor smooth muscle. To examine the composition of biopsy samples we used RT-qPCR to determine the expression of specific gene markers of each layer: Desmin (*DES*) for smooth muscle, Uroplakin (*UPK2*) for urothelium, and Vimentin (*VIM*) for myofibroblast/stromal cells. Primers were validated as shown in Supplementary Figure 1 and detailed in the methods.

As shown in Figure 1a, expression of the smooth muscle marker *DES* was decreased in rOAB biopsies compared to controls (p=0.01, n=4 control and n=26 OAB samples). In contrast, expression of the myofibroblast/stromal marker *VIM* and the urothelium marker *UPK2* was not different between sample types. A high degree of variation was observed in biopsy composition for some markers when using fold change analysis, but it was evident from the relative Cq values of the different markers within each biopsy that some samples predominantly expressed one or two markers, while others expressed equal levels of all three. Samples were therefore grouped into six different subgroups (*DES*^hi^, *VIM*^hi^, *UPK2*^hi^, *DES&VIM*^hi^, *VIM&UPK2*^hi^, and EVEN) based on the relative expression of tissue markers. Strikingly, most control samples (75%) but only 31% of rOAB samples were classified as *DES*^hi^ (Figure 1b). Together these data suggest that relative smooth muscle content is reduced in the bladder of rOAB patients.

**Figure 1.**
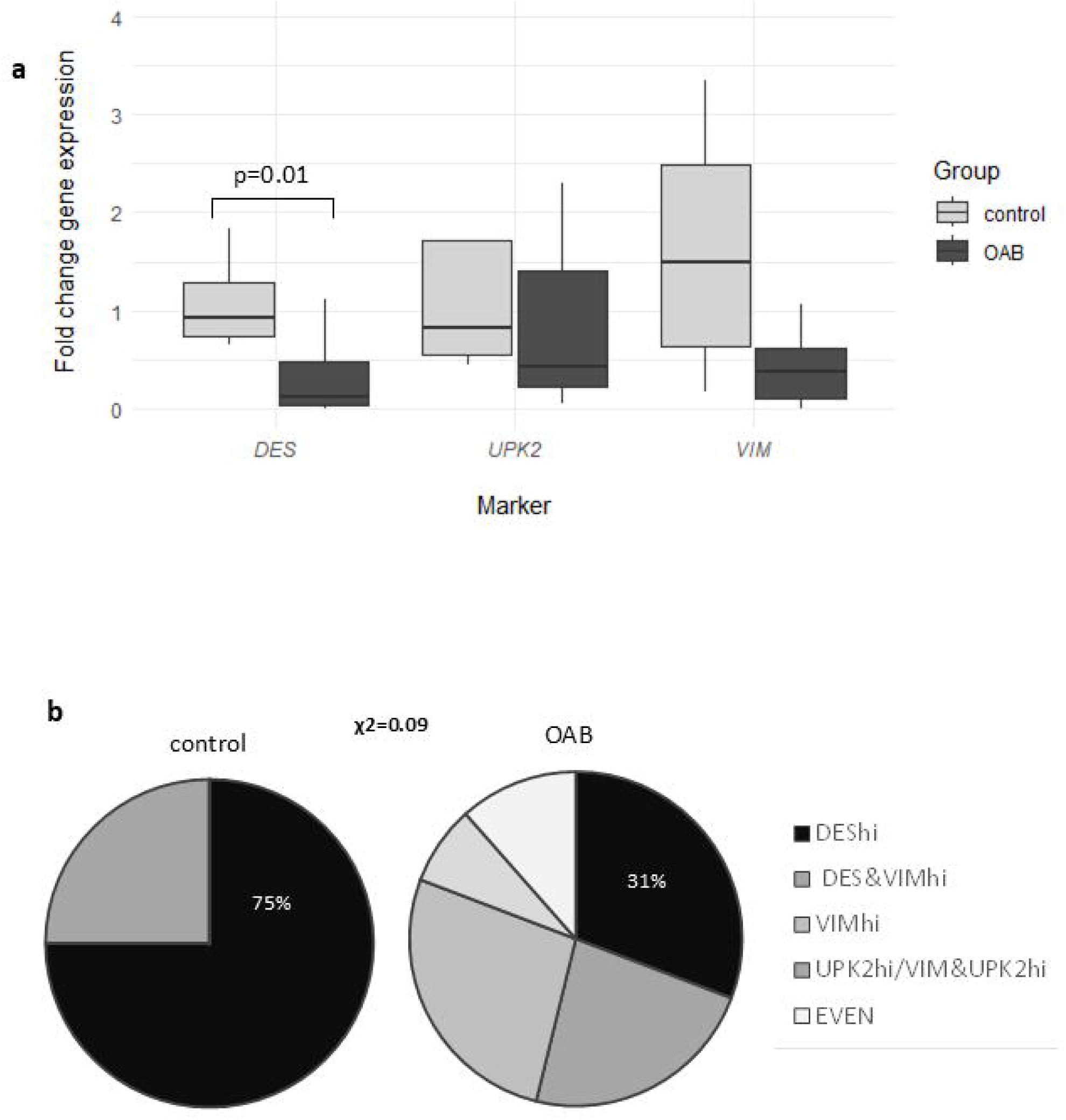
Analysis of cellular composition in control and OAB biopsies. Expression of markers for smooth muscle (*DES*), stroma (*VIM*) and urothelium (*UPK2*) was quantified by RT-qPCR to determine the relative contribution of each cell type to biopsy composition. In (a) fold change gene expression was determined using the ΔΔCq method and a normalisation factor derived from 3 reference genes, with fold change calculated relative to the mean control value. Statistical analysis was carried out in R studio using a Mann-Whitney U test due to small control sample size (n=4 control and n=26 OAB samples; RNA was not available for 2 control and 7 OAB patients). Graph shows a box and whisker plot. In (b), samples were categorised into the indicated subgroups based on predominant marker/s expression and presented as a pie chart. A χ2 test was performed to determine statistical significance of the difference in the distribution of DES^hi^/other sub-groups between control and OAB samples. The categorisation is described fully in the supplementary methods.

### MGF expression in rOAB tissue

MGF expression has been extensively studied in skeletal muscle tissue and was only previously investigated in the bladder by Cortes *et al* in 2011^17^. As shown in Figure 2a, MGF protein expression was detected in bladder biopsies by immunohistochemistry, where a diffuse stain was observed throughout the biopsy tissue. MGF protein expression was confirmed by western blot analysis of biopsy lysates using an antibody specific to the MGF-E peptide that recognizes both pro- and mature MGF, but not IGF-I. As shown in Figure 2b, bands of around 15 kDa were observed, consistent with previous reports^18–20^ and likely representing the MGF precursor protein. Quantification of MGF protein expression by western blot showed that mean MGF expression is similar in control and rOAB biopsies (Figure 2c). Analysis of MGF expression in each of the main sub-groups identified in Figure 1 demonstrates that MGF expression is highest in *DES*^hi^ rOAB samples, but MGF expression in different sub-groups is not different between control and rOAB samples (Figure 2d). In summary, therefore, no significant difference in MGF expression was detected between control and rOAB biopsies.

**Figure 2.**
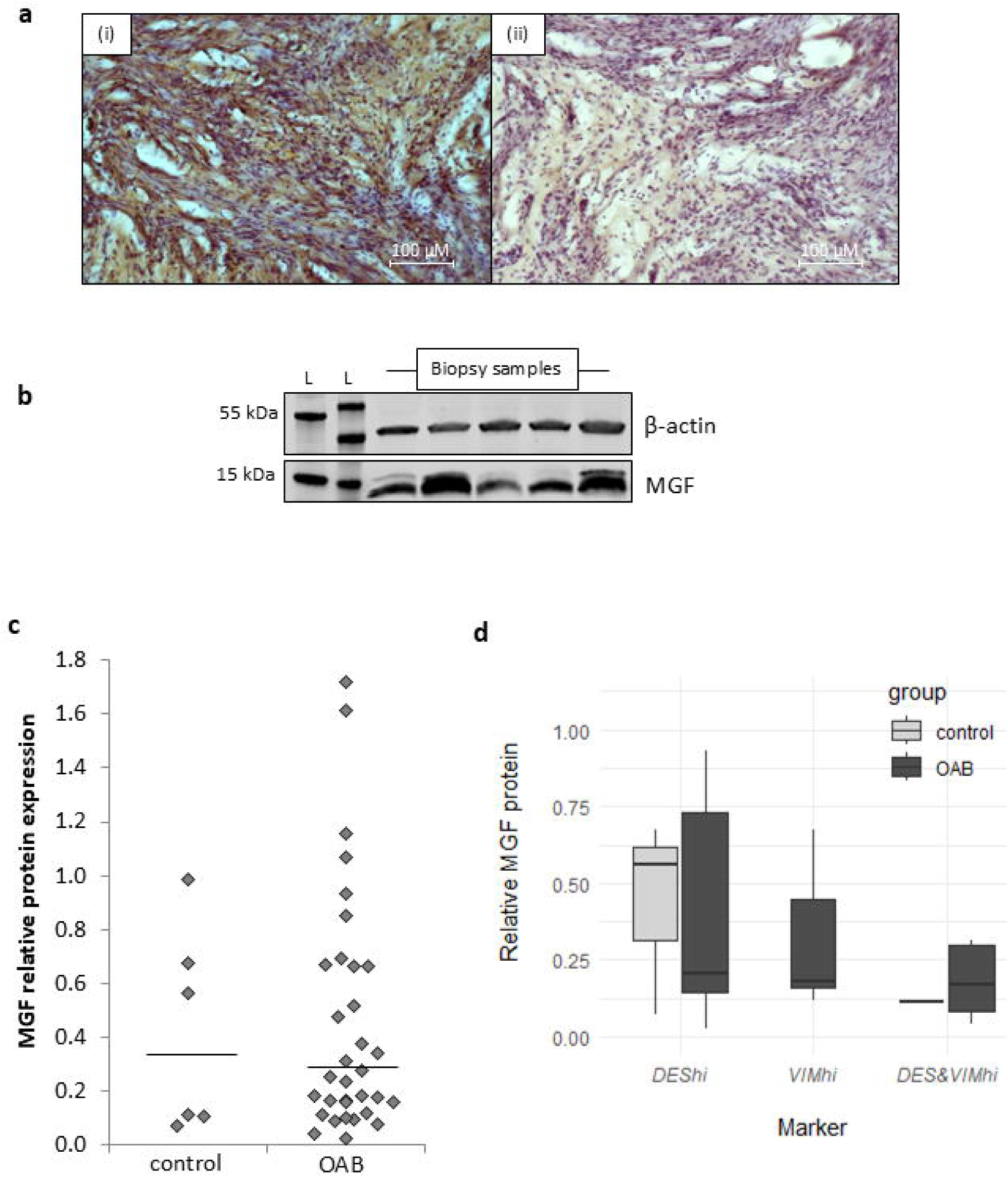
MGF expression in OAB tissue. An antibody specific to the MGF E-peptide was used to detect expression of MGF protein by immunohistochemistry staining (a) and western blot (b-d) analysis of biopsy sections. In (a) 5µm sections of formalin-fixed OAB bladder biopsies were stained with anti-MGF (i) or secondary antibody only control (ii) and counterstained with haematoxylin. In (b-d) 30µg biopsy lysates were analysed by western blot for detection of MGF (∼15 kDa) and β-actin (42 kDa) as a loading control. L=size ladder. A representative blot is shown in (b) and a scatter plot showing quantification of MGF band intensity is shown in (c). The bar indicates median relative MGF expression in control and OAB biopsies: 0.34 (n=6) and 0.25 (n=33), respectively (p=0.8). In (d), a box and whisker plot of relative MGF expression in control/OAB biopsies is shown for the three main sub-groups identified in Figure 1: *DES*^hi^ (n=3/8; p=1.0), *VIM*^hi^ (n=0/7), *DES&VIM^hi^* (n=1/6). No *VIM*^hi^ samples were seen in controls. Statistical analysis, where sample size allowed, was performed using Mann-Whitney U test.

### Analysis of correlation between MGF protein expression and clinical parameters of OAB severity

To investigate the relationship between MGF expression and clinical severity of OAB, we looked at the urodynamic parameter maximum cystometric capacity (MCC) and we looked at validated screening questionnaire scores from OAB-V8 and BSAQ, used clinically in rOAB patients to establish the perseverance of symptoms following failed conservative therapy. A higher MCC score indicates a greater bladder capacity and in principle less clinically severe OAB, whereas a high questionnaire score indicates more clinically severe OAB. As shown in Figure 3, there was no significant correlation between MCC score and MGF expression (R=0.29, p=0.18 n=23; clinical parameters were not available for all patients). Similarly, no correlation was detected between MGF expression and either BSAQ or OABV8 self-assessment scores (BSAQ: R=0.29, p=0.14, n=27; OABV8: R=0.18, p=0.37, n=27). We therefore tested the correlation between questionnaire scores and MCC however, as shown in Supplementary Figure 2, although positive correlation was seen between BSAQ and OABV8 scores (R=0.72, p=0.00002, n=27), no correlation was observed between MCC score and self-assessments, reflecting the limitations of current clinical tools in measuring OAB severity.

**Figure 3.**
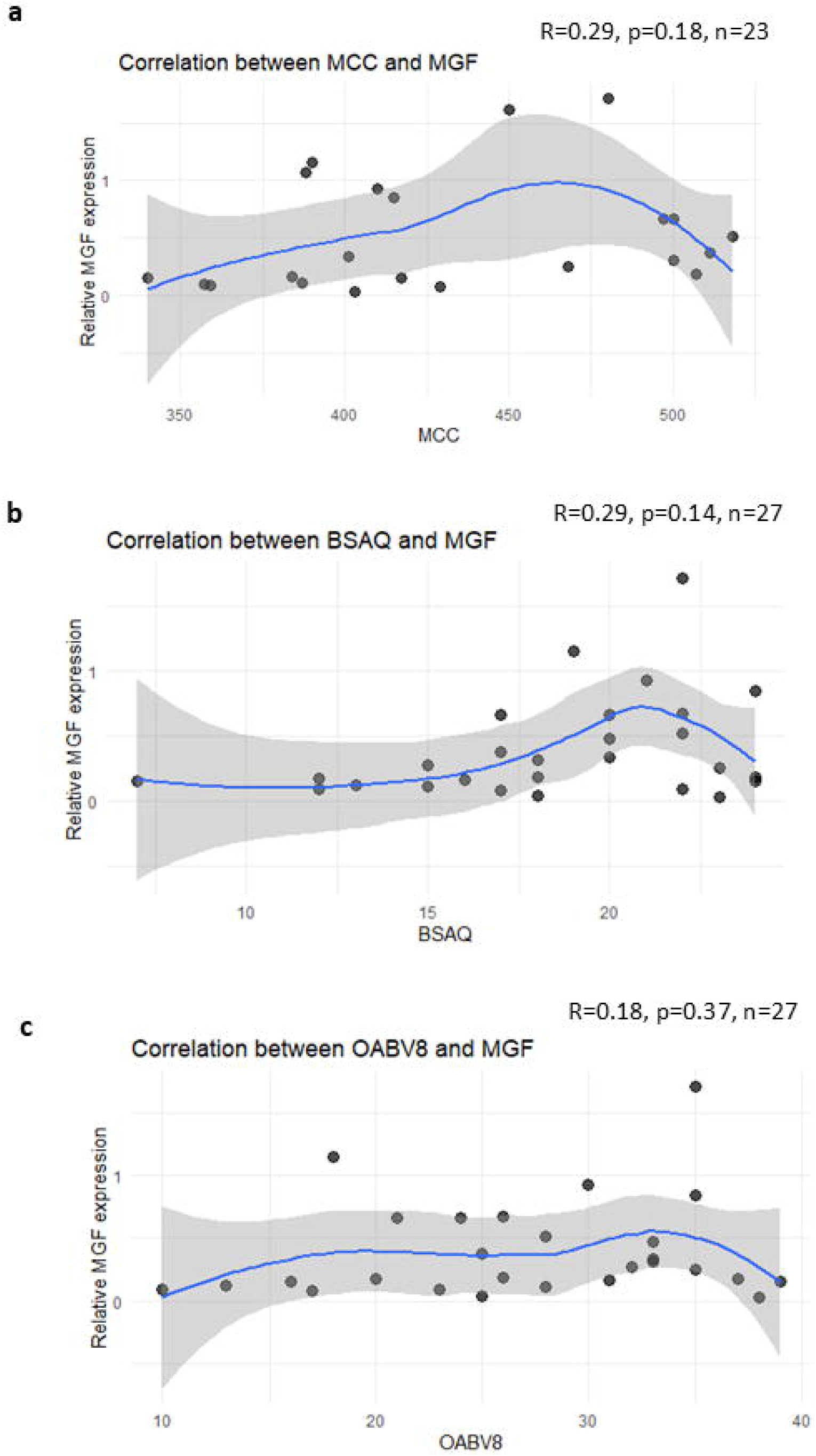
Analysis of correlation between MGF protein expression and clinical parameters of OAB severity. Spearman’s correlation was used to determine the correlation between biopsy MGF expression and (a) the maximum cystometric capacity (MCC) in OAB patients, and between MGF expression and patient self-assessment scores for two-types of questionnaire: BSAQ (b) and OAB-V8 (c). In (a) one data point where MCC<250 is not shown in the graph for visual clarity.

When we looked at the difference in MGF expression in women of pre- and post-menopausal age there was a marginal increase in MGF expression in women over the age of 50 with OAB compared to women with OAB who were less than 50 years of age, though this difference was not statistically significant (Supplementary Figure 3). No difference in MGF expression was observed in the two age groups in control patients, although it should be noted that patient numbers in these groups were small. Together this suggests that there is limited evidence for an association between hormonal changes associated with menopausal age and MGF expression in the bladder.

### MGF promotes smooth muscle cell proliferation *in vitro*

To test whether MGF can promote bladder smooth muscle proliferation *in vitro*, single cell suspensions from murine bladder were prepared and cultured for 72-hours in the presence of varying concentrations of MGF. Proliferation was determined by EdU assay, co-staining with an antibody to smooth muscle myosin heavy chain (SMMHC) to identify smooth muscle cells within the mixed population. Staining indicated that most cells were SMMHC positive smooth muscle cells (Figure 4a). As shown in Figure 4b, addition of increasing concentrations of MGF induced increasing levels of smooth muscle cell proliferation. At the highest concentration used (100 ng/ml), MGF peptide induced a more than three-fold increase in the percentage of proliferating nuclei compared to untreated controls (4.4% ± 0.02 in control, 14.1% ± 0.03 with 100 ng/ml of MGF, p=0.008). This supports the hypothesis that MGF may promote bladder smooth muscle regeneration.

**Figure 4.**
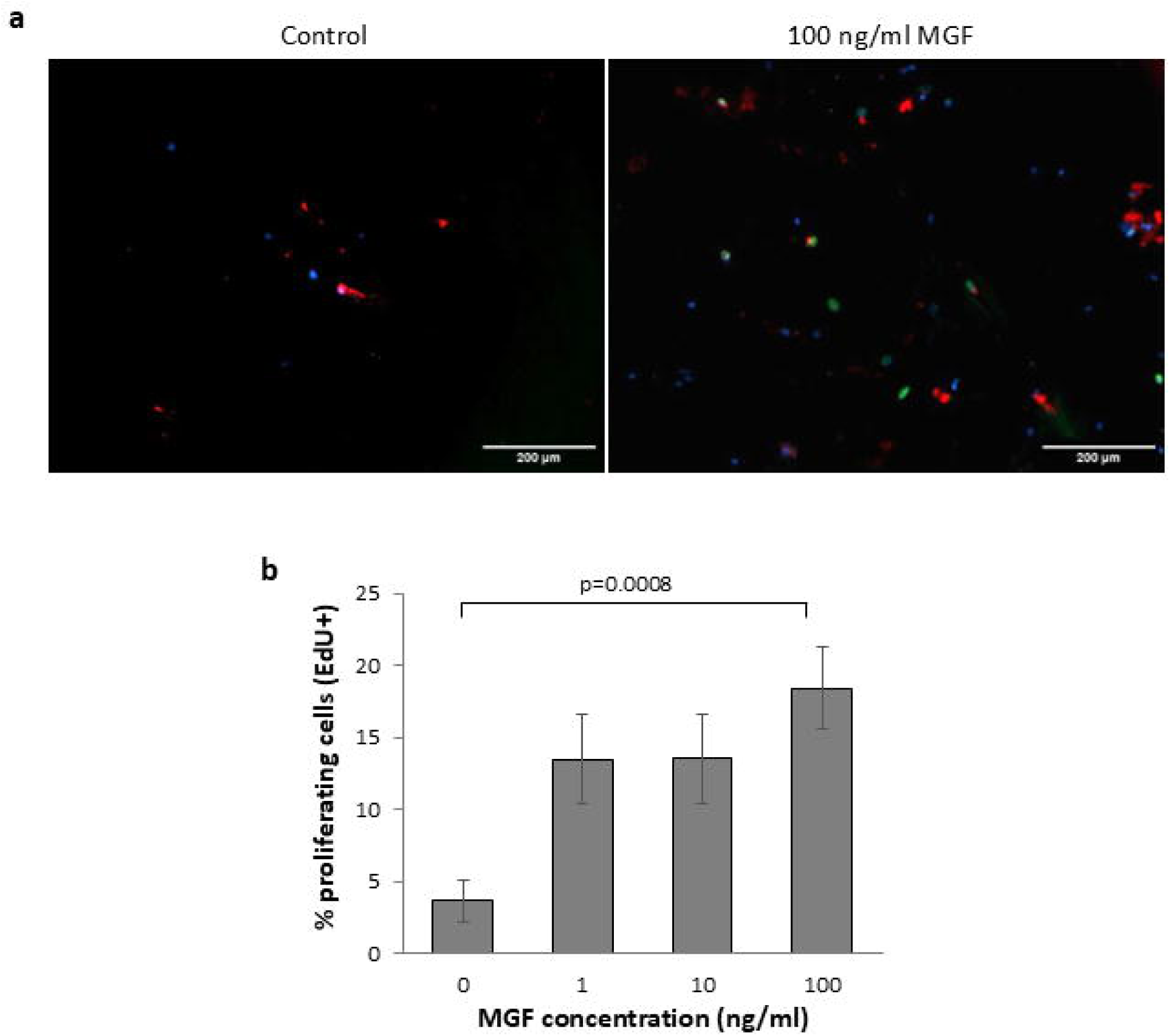
Effect of MGF on primary murine bladder cells proliferation. The indicated concentration of synthetic MGF peptide was added to primary cultures of murine bladder cells for 72 hours, with EdU added for the final 48 hours. Cells were stained with an anti-SMMHC antibody (red) to identify smooth muscle cells, and EdU was visualised by Click-iT® detection (green); all cells were labelled using DAPI nuclear stain (blue). Images were taken at 4X magnification using an EVOS-imaging station, with additional digital zoom. Representative images are shown in (a), and the graph in (b) shows mean %EdU+ proliferating cells/image +/- SEM (n=11-18 images, from 3-5 wells). Statistical analysis was performed using one-way ANOVA test followed by post-hoc t-test.

## Discussion

Our data shows that MGF is expressed in the bladder and can be detected at the protein level. We have further shown that MGF promotes cellular proliferation in bladder smooth muscle, consistent with studies in myoblast^10,12^ and human skeletal muscle cells^21^. It is interesting to speculate on the effect of MGF in the complex bladder physiology, since MGF has been shown to have regenerative properties in both neuronal and muscle tissue. The potential effect of MGF in the lamina propria is of particular interest since it can be thought of as the functional centre of the bladder wall, consisting of extracellular matrix (ECM), progenitor cells, interstitial cells, and efferent nerve endings^22^, all upon which MGF may exert its regenerative action.

Increased hypoxic conditions in the bladder have been shown to alter cell metabolism, promoting angiogenesis, and apoptosis^23^. This in turn increases expression of growth factors such as nerve growth factor (NGF), tumour growth factor-B (TGF-B) (which promotes remodelling of the ECM and fibrosis), vascular endothelial growth factor (VEGF)^24^, but also potentially MGF as seen in neuronal tissue^15^. This could imply that a reduced bladder perfusion, seen as causative factor in OAB^25^, could be prevented by a healthy MGF pool provision due to its anti-apoptotic properties^26^.

Studies have revealed that mechanically stressed bladder smooth muscle cells are acutely receptive to their biomechanical environment and cyclical deformation induces connective tissue synthesis^27,28^. Although the mechanisms underlying these effects have yet to be determined, the basis of mechano-transduction, the process by which muscle cells detect mechanical force and convert this into biochemical signalling, is beginning to be better understood^29^. As described by Comperat *et al*, bladder histology from patients with OAB show a loss of smooth muscle with replacement by patches of fibrosis and tissue high in collagen. This process seems to revert following treatment with botulinum toxin A injections into the bladder wall which promote further relaxation and hence distension of the bladder wall^27^. In line with this, we have found that on analysis of bladder biopsies, the level of expression of the smooth muscle marker *DES*^hi^ (detecting smooth muscle cells of the detrusor muscle) was reduced in our rOAB study group compared with our control group. Increased deposition of connective tissue between muscle fascicles is a well-characterised pathological feature of OAB^30^. However, it should be noted that we cannot rule out the effect of sampling variation as biopsy sample depth is not controlled.

In summary, our study found similar expression of MGF in control and rOAB bladder biopsies, and no correlation between MGF and symptom severity. This suggests that a change in MGF expression is unlikely to be involved in the development of OAB. However, although preliminary, our study demonstrates that treatment with MGF induces bladder cell proliferation *in vitro*, suggesting the therapeutic potential of MGF. It is known that mechanical distension triggers MGF activity in skeletal muscle^10^ and it is not unreasonable to speculate that this process may be replicated following therapies that aim to facilitate bladder distension. It will be of interest in future studies to determine whether individual conditions that favour splicing to produce MGF protein are associated with better response to conservative OAB therapy. Further research is therefore needed to establish the sequence of events between aetiology, symptomatology and therapy.

## Data Availability

All data produced in the present study are available upon reasonable request to the authors

## Dedicatory

We would like to dedicate this paper to Professor Linda Cardozo MD FRCOG OBE from Kings College Hospital for her support during the length of this project, who passed away unexpectedly before completion of the manuscript.

## Statement of contribution

ES designed the experiments, carried out the majority of experimental work, analysed and interpreted data, drafted the figures and prepared the manuscript; BL consented patients, collected biopsy samples and clinical data, and contributed to clinical sections of the manuscript; FM-Y contributed data to Figure 1; AMu contributed to Figure 4; AMa was involved in assay development; RB consented patients, collected biopsy samples and clinical data; FEM was involved in project supervision and contributed to discussions on data analysis and interpretation; NJH designed experiments, supervised the project, contributed to data analysis and interpretation, prepared the figures and co-wrote the manuscript; EC conceived the project, obtained ethical permissions and funding, recruited participants and collected biopsy samples and edited the manuscript. All authors have read and agreed to publication of this manuscript.

