## Supplementary methods and figures for "Expression of mechano-growth factor (MGF) in refractory overactive bladder"

**RNA isolation and cDNA synthesis**

RNA was quantified using a BioDrop Spectrophotometer (Fisher), and 260/280 ratio used as an indication of sample purity. Total mRNA (40 ng/reaction) was reverse transcribed using a RevertAid First Strand cDNA Synthesis Kit (Fisher) with oligo(dT) primer. 'No reverse transcriptase' control (NRT) and 'no template control' (NTC) reactions were used as negative controls for the cDNA synthesis. The reaction was performed in Applied Biosystems Veriti 96 well thermal cycler, using the following thermal profile: 42° C for 60 ', 70° C for 5 '.

**RT-qPCR and sub-group analysis**

RT-qPCR was performed using Maxima SYBR Green Master Mix (Fisher), 4.5 ng RNA equivalent cDNA and appropriate forward and reverse primers (see Supplementary Figure 1g for primer sequences) in a final volume of 5µl. Amplification of NRT, NTC and $dH_20$ negative controls was also performed. RT-qPCR amplification was performed in a Techne Prime Pro 48 Real-time qPCR machine, using the following thermal profile: 95° C for 10'; 40 cycles of 95° C for 10", 55° C for 30" and 72° C for 15'', followed by one cycle of 95° C for 15", 55° C for 15" and 95° C for 15". Melt curve analysis and agarose gel electrophoresis were used to confirm amplification of a single PCR product. Primer efficiency was determined by amplification of serially diluted templates. Relative quantification was performed using the ΔΔCq method[1].
Samples were tested for expression of desmin (*DES*), vimentin (*VIM*), uroplakin 2 (*UPK2*) and β2-microglobulin (*B2M*). Three different reference genes were used (*GAPDH, B2M, ACTB*) and a normalisation factor based on the arithmetic mean of the Cq value of all three genes was used to control for loading variation. RNA was not available for all samples therefore RT-qPCR analysis was only possible on a subset of the cohort.
For the sub-group analysis, relative inverted Cq (RI-Cq) values were first calculated by subtracting each Cq value from 41 (the maximum cycle number + 1) and then calculating as a percentage relative to the total inverted Cq value for all 3 tissue marker genes in that biopsy. This provides a rough estimation of the relative contribution of each cell type within a biopsy. A sample was designated as $DES^{hi}$ where *DES* was the major component in the biopsy *ie* if the RI-Cq value for *DES* was higher than *VIM* and *UPK2* by >3. Similar designations were used for $VIM^{hi}$ and $UPK2^{hi}$ groups. Where two genes were contributing equally to the sample the designation was eg $DES\&VIM^{hi}$, and where the difference between all three genes was <3 the sample was designated as 'EVEN'.

**Immunohistochemistry**

Human OAB bladder samples were fixed in 10% neutral buffered formalin for 24 h, embedded in paraffin, and sectioned at 5-µm thickness onto poly-L-lysine coated slides. Antigen retrieval was performed by boiling in 0.01M citrate buffer for 10 mins. Sections were stained using an anti-MGF E-peptide antibody (Merck Life Sciences #07-2108, rabbit polyclonal, 1/50 dilution) and HRP-tagged secondary (Vector Laboratories, BA-1100, 1/200 dilution), or secondary only control staining. Detection was performed using the Vector ABC kit with 3,3′-Diaminobenzidine (DAB) as substrate. Sections were counterstained with haematoxylin and imaged using a Nikon H550L microscope and 20X objective.

**Western blot**

Total protein concentration was quantified using the Bicinchoninic Acid (BCA) assay (Pierce), and 30 µg protein/well was run for ~1.5 hrs on a Mini-PROTEAN TGX Precast Protein Gel (Biorad) before transfer to nitrocellulose using a Trans-Blot Turbo (Biorad). Membranes were blocked in Odyssey TBS blocking buffer (Li-Cor) for 1 h, washed with Tris-buffered saline with Tween 20 (TBST) and incubated with diluted anti-MGF (Sigma-Aldrich #07-2108, rabbit polyclonal raised against the E-peptide of MGF; 1/1,000 dilution) and anti-β-actin (mouse monoclonal #8224, Abcam, 1/1,000 dilution) primary antibodies at 4°C for 24 h. Blots were washed and incubated with 1/15,000 of near-infrared (NIR)-labelled secondary antibodies for 1hr at RT (anti-mouse-IR680LT (red) and anti-rabbit-IR800CW (green), both from Li-Cor). After washing, blots were visualised using the Li-Cor Odyssey CLx NIR imager and signal quantified using Li-Cor Image Studio Lite V.5.2 software. MGF band intensity was normalized to β-actin loading control.

**Isolation and culture of murine bladder smooth muscle cells**

Murine bladder was isolated from adult outbred ICR mice aged 8-12 weeks (Harlan, Huntingdon UK) following cervical dislocation, carried out at Kings College London (establishment licence: X24D82DFF; project licence: PBCFBE464) in accordance with the U.K. Home Office Animals (Scientific Procedures) Act 1986 with 2012 amendments. Tissue was washed thoroughly in D-PBS containing antibiotics (pen/strep) and anti-mycotic agent (Fungizone, Gibco). A single cell solution containing smooth muscle cells was prepared based on a modified version of the protocol by Pokrywczynsja *et al.*[2]. Briefly, bladders from 3 animals were minced using sterile scalpels and scissors and incubated at 37°C for 1 hr in digestion solution (1 ml Advanced DMEM/F-12 supplemented with antibiotics and anti-mycotic, 2 mg/ml Collagenase D and Dispase II). Enzymes were then blocked by adding an equal amount of medium and the solution was passed through a 100 µm cell strainer to remove undigested tissue. The suspension was then centrifuged for 5 mins at 1500 X g, the supernatant discarded, and the cell pellet resuspended in smooth muscle (SM) culture medium supplemented with 5% smooth muscle growth supplements (SMGS, Gibco). The cells were seeded at 0.01 x $10^6$ cells/well in a 96-well plate and fresh medium added each day.

MGF peptide with the sequence Tyr-Gln-Pro-Pro-Ser-Thr-Asn-Lys-Asn-Thr-Lys-Ser-Gln-D-Arg-D-Arg-Lys-Gly-Ser-Thr-Phe-Glu-Glu-His-Lys[3] was commercially synthesized (Genscript) and used to treat the bladder cell cultures for 72 hours at the indicated concentrations, after first allowing the cells to adhere overnight.

**EdU staining assay and cell viability assay.**

Cell proliferation was assessed by EdU (5-ethynyl-2'-deoxyuridine) incorporation into primary murine bladder cells (EdU-Click 488, Sigma-Aldrich), following the manufacturer's guidelines, for the last 48 hrs of 72 hr culture. EdU is incorporated into DNA during active DNA synthesis. Cells were fixed in 4% PFA, permeabilised with 0.4% Triton-X100/PBS, washed and blocked with 0.05M glycine buffer. Cells were then stained with rabbit anti-smooth muscle myosin heavy chain 11 (SMMHC) primary antibody (Abcam, #53219, 1/100) for 1 hr at RT, to identify smooth muscle cells[4]. Cells were then incubated in secondary antibody for 1 hr at RT (IgG (H+L) Highly Cross-Adsorbed Donkey anti-Rabbit, Alexa Fluor 555, Invitrogen; 1/200), and counterstained with DAPI. Images were taken using an EVOS Imaging Station (4X objective). Quantification of total and proliferating cells, labelled with Alexa 488, was performed using the cell count function and manually, respectively, of ImageJ software[5].

### Supplementary Figures

### Supplementary Figure 1

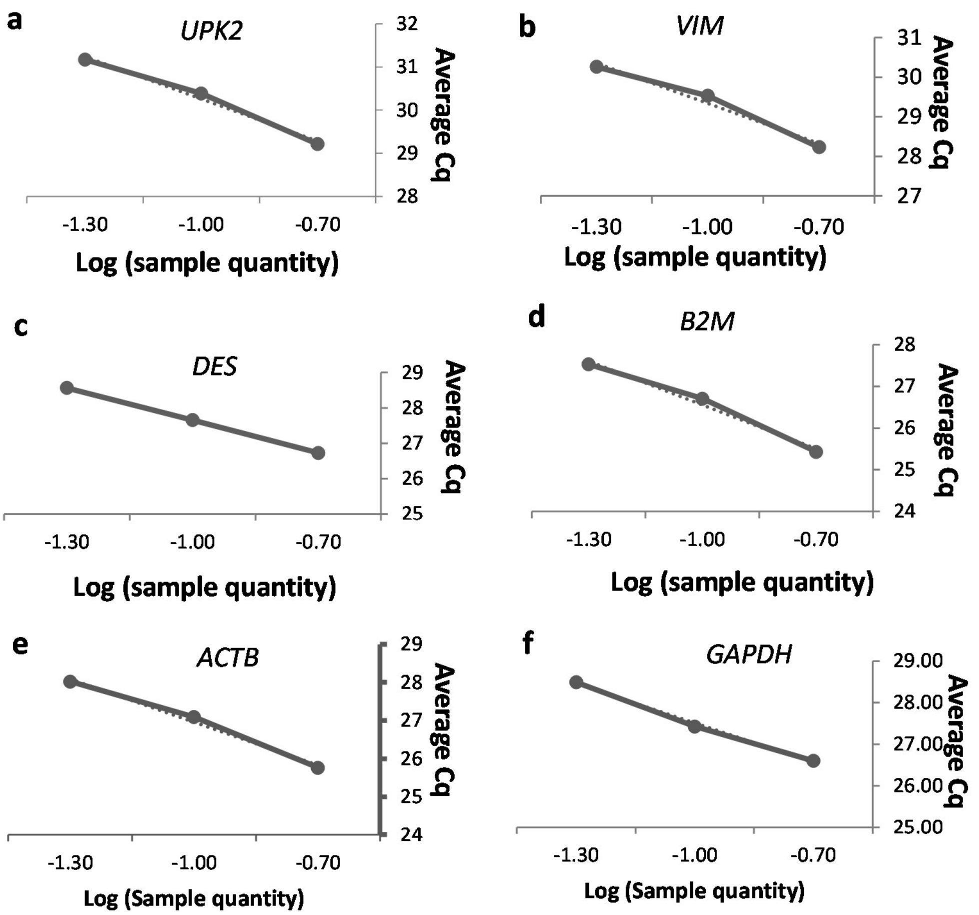


g

| | *DES* | *VIM* | *UPK2* | *B2M* | *ACTB* | *GAPDH* |
|---|---|---|---|---|---|---|
| **Slope** | -3.05 | -3.37 | -3.25 | -3.49 | -3.8 | -3.14 |
| **RSQ** | 1.00 | 0.97 | 0.99 | 0.98 | 0.99 | 1.00 |
| **Primer Efficiency (%)** | 112.9 | 98.1 | 103.0 | 93.6 | 84.7 | 108.4 |

h

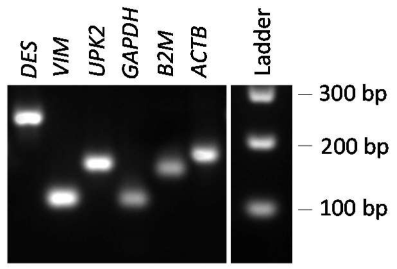


i

| Name | Gene ID | For primer (5'->3') | Rev primer (5'->3') | Product size (bp) |
|---|---|---|---|---|
| ***DES*** | 1674 | ACCATCGCGGCTAAGAACAT | TCACTGGCAAATCGGTCCTC | 230 |
| ***VIM*** | 7431 | GTTGACAATGCGTCTCTGGC | GCAGCTCCTGGATTTCCTCT | 109 |
| ***UPK2*** | 7379 | CTCCCTCGAAGGAACATGGAA | AGACCTCCTTACTTGCGGGA | 152 |
| ***B2M*** | 567 | TGTCTTTCAGCAAGGACTGGT | TGCTTACATGTCTCGATCCCAC | 143 |
| ***GAPDH*** | 2597 | CCTCCTGTTCGACAGTCAGC | ACGACCAAATCCGTTGACTCC | 105 |
| ***ACTB*** | 60 | GCCGCCAGCTCACCA | ATCCTTCTGACCCATGCCCA | 167 |

**Supplementary Figure 2**

R=0.72, p=0.00002, n=27

**a**

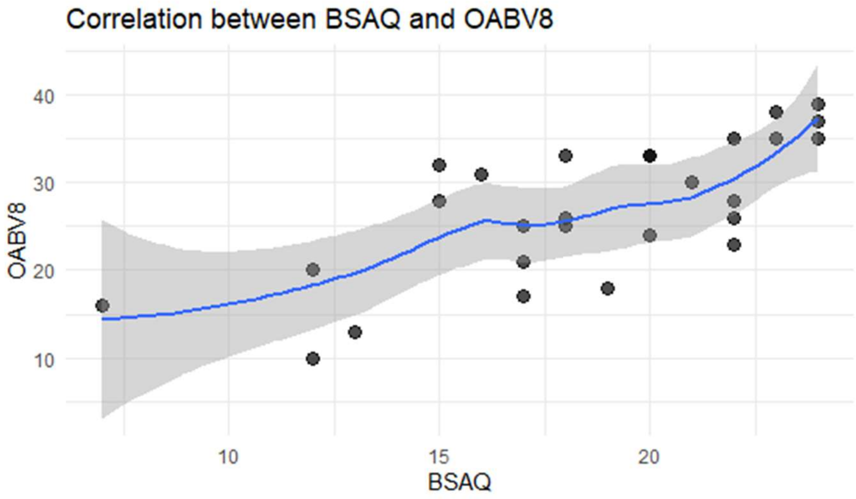


**b**

R=0.08, p=0.73, n=20

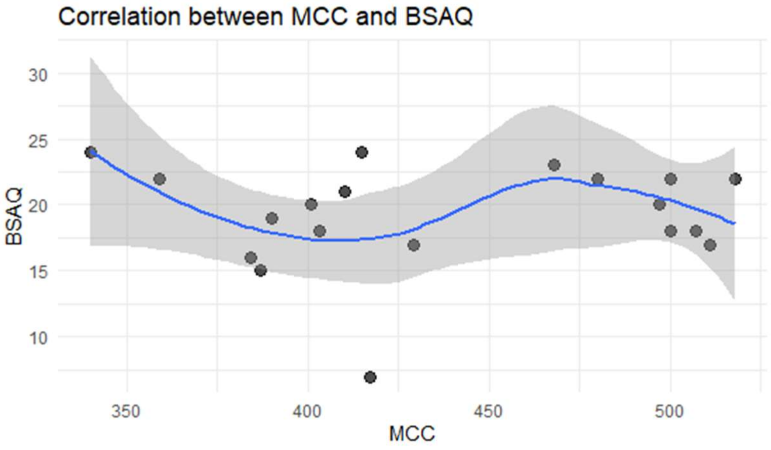


R=0.03, p=0.91, n=20

**c**

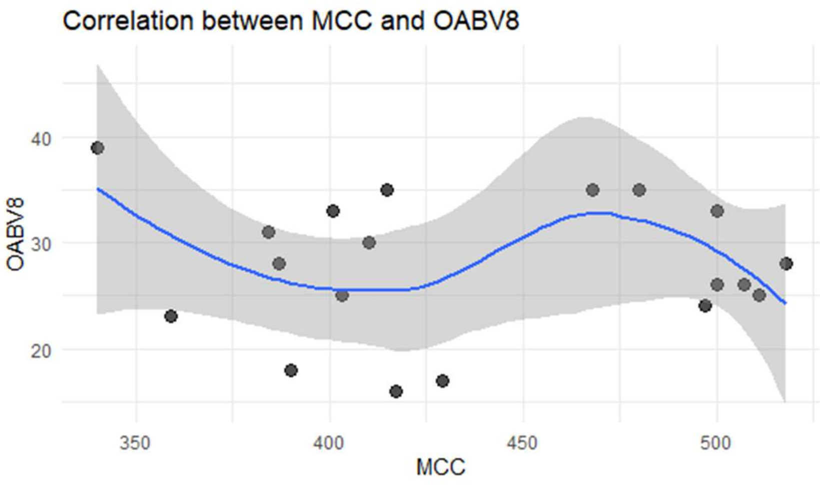

**Supplementary Figure 3**

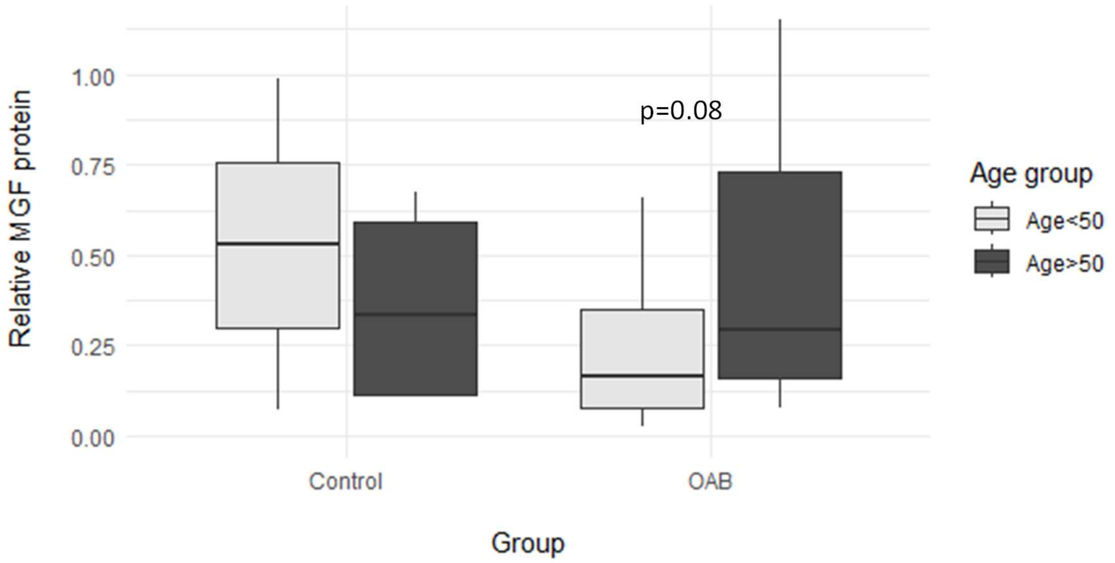


### Supplementary Figure legends

**Supplementary Figure 1: Primer validation for RT-qPCR.** The amplification efficiency of *UPKN* (a), *VIM* (b), *DES* (c), *B2M* (d), *ACTB* (e) and *GAPDH* (f) primers used for RT-qPCR biopsy analysis was determined by serial template dilution and plotting Cq values against log (sample quantity). The slope of the linear best fit was used to calculate primer efficiency (g). In (h), RT-PCR products amplified using the indicated primers on a biopsy sample were analysed by agarose gel electrophoresis. Single bands of the expected size were observed with all primers used (*DES, VIM, UPK2, GAPDH, B2M* and *ACTB*). The expected PCR product sizes in addition to primer sequences are shown in (i).

**Supplementary Figure 2: Analysis of correlation between clinical parameters of OAB severity.** Spearman's correlation was used to determine the relationship between BSAQ and OABV8 clinical questionnaire scores in OAB patients (a). Correlation was also tested between MCC and patients' scores from BSAQ (b) and OAB-V8 (c) questionnaires. In (b) and (c) one data point where MCC<250 is not shown in the graph for visual clarity.

**Supplementary Figure 3: Analysis of MGF expression in patients of pre- and post-menopausal age**. A box and whisker plot of MGF expression is shown for both control and OAB patients designated either pre- or post-menopausal age (≤50 or >50 yrs, respectively). Mann-Whitney U test was used to determine statistical significance (n values for patients ≤50 and >50 yrs are 2/4 for controls, 8/24 for OAB, respectively; age information was not available for one patient).
